# Clinical Features of COVID-19-Related Liver Damage

**DOI:** 10.1101/2020.02.26.20026971

**Authors:** Zhenyu Fan, Liping Chen, Jun Li, Cheng Tian, Yajun Zhang, Shaoping Huang, Zhanju Liu, Jilin Cheng

## Abstract

**BACKGROUND:** A recent outbreak of SARS-CoV-2 infection occurs mainly in China, with rapidly increasing the number of cases (namely COVID-19). Abnormal liver functions are frequently present in these patients, here we aimed to clarify the clinical features of COVID-19-related liver damage to provide some references for the clinical treatment.

**METHODS:** In this retrospective, single-center study, we included all confirmed COVID-19 cases in Shanghai Public Health Clinical Center from January 20 to January 31, 2020. The outcomes were followed up until February 19, 2020. A total of 148 cases were analyzed for clinical features, laboratory parameters (including liver function tests), medications and the length of stay.

**FINDINGS:** Of 148 confirmed SARS-CoV-2-infected patients, 49.3% were females and 50.7% were males. The median age was 50.5 years (interquartile range, 36-64). Patients had clinical manifestations of fever (70.1%), cough (45.3%), expectoration (26.7%) at admission. 75 patients (50.7%) showed abnormal liver functions at admission. Patients (n = 75) who had elevated liver function index were more likely to have a moderate-high degree fever (44% vs 27.4%; p = 0.035) and significantly present in male patients (62.67% vs 38.36%; p = 0.005). The numbers of CD4^+^ and CD8^+^ T cells were significantly lower in abnormal liver function group than those in normal liver function group. There was no statistical difference in prehospital medications between normal and abnormal liver function groups, while the utilization rate of lopinavir/ritonavir after admission was significantly higher in patients with emerging liver injury than that in patients with normal liver functions. Importantly, the emerging abnormal liver functions after admission caused a prolonged length of stay

**INTERPRETATION:** SARS-CoV-2 may cause the liver function damage and the Lopinavir/ritonavir should be applied carefully for the treatment of COVID-19.

**FUNDING:** Shanghai Science and Technology Commission Fund Project and National Science and Technology Major Project

## Introduction

Since December 2019, a novel coronavirus was identified as the pathogen to cause pneumonia in Wuhan, China, which was temporarily named as 2019-nCoV by WHO.^2,3^ On 11 February 2020, based on the phylogeny, taxonomy and established practice, 2019-nCoV was officially named as SARS-CoV-2, ^4^ and the disease caused by SARS-CoV-2 was named as COVID-19.^5^ Evidence has proven that SARS-CoV-2 can be transmitted from person to person through respiratory droplets and close contact, posing a huge challenge on the public health.^6^ Up to now, there were more than seventy thousand confirmed cases and two thousand death case in 30 countries around the world.

The main manifestations of SARS-CoV-2 infection include fever, dry cough, weakness, and breathing difficulty. Abnormal liver functions were frequently reported extrapulmonary clinical feature and almost one half of patients experienced different degrees of liver function damage. ^8,9^.According to the recent study of single-cell RNA-seq, angiotensin converting enzyme(ACE)2 was highly expressed not only in type II alveolar epithelial cells, but also in bile duct cells.^10^ Importantly, recent study confirmed that the cell entry receptor of 2019-nCoV, like SARS-CoV, is ACE2 receptor.^11^ All these findings suggest that SARS-CoV-2 may infect the bile duct cells and cause the abnormal liver function in these patients. However, alkaline phosphatase (ALP), the bile duct injury related index, was not specific in COVID-19 patients.^8,9^.A recent pathological finding reported that moderate microvascular steatosis and mild lobular and portal activity were present in liver biopsy specimens, indicating that the liver injury could be caused by either SARS-CoV-2 infection or drug-induced liver injury.^12^ Currently, however, there is no any clinical research to analyze whether abnormal liver function in COVID-19 patients is due to the drugs used or not. Given the high contagious and pathogenic capacities of SARS-CoV-2 and high incidence of liver damage, an analysis with liver function in COVID patients is urgently warranted. In this study, we retrospectively investigated the liver function change in SARS-CoV-2-infected patients from a single center study in Shanghai, China, and compared the clinical features, medications and length of stay of COVID-19 patients with/without liver damage. The purpose of this study is to clarify the clinical features of COVID-19-related liver damage, evaluate the association between current medications and liver damage, and therefore provide a reference for clinical treatment of patients with COVID-19.

## Methods

### Study design and participants

From January 20, 2020 to January 31, 2020, a total of 148 consecutive patients were admitted and treated in the Shanghai Public Health Clinical Center affiliated to Fudan University (the designated hospital in the Shanghai area), all of which were confirmed cases of COVID-19. The clinical criteria of diagnosis and discharge refer to the standards for “Diagnosis and Treatment Scheme of New Coronavirus Infected Pneumonia” (trial version 6).^13^All cases had a history of related epidemiology, most of them had clinical manifestations, such as fever or respiratory symptoms. All patients were diagnosed after examination of SARS-CoV-2 RNA by RT-PCR and chest CT scanning. By February 19, 93 of the 148 cases were discharged from the hospital, and 1 died during this period. The average length of hospitalization of discharged cases was 12 days (6-21 days). This study was approved by the Ethics Committee of the Shanghai Public Health Clinical Center (2019-S047-02, Review date: Jan 13, 2020) and was exempted from the need for informed consent from patients.

### Data collection

The medical records of 148 patients were collected and analyzed by the research team from the Department of Gastroenterology and Hepatology, Shanghai Public Health Clinical Center, Fudan University. Epidemiological, clinical, laboratory characteristics and treatment and outcomes data were acquired by the hospitalization management system.

### Laboratory examination

The sera were harvested from all confirmed patients after an overnight fast. All laboratory data were obtained on the same day as the sera were extracted. Laboratory examination was conducted every three days. Peripheral leukocyte count, lymphocyte absolute value, erythrocyte sedimentation rate (ESR), procalcitonin (PCT), liver functions including alanine aminotransferase (ALT, 40 U/L), aspartate aminotransferase (AST, 13-35 U/L), lactate dehydrogenase (LDH, 109-245 U/L), gamma-glutamyltransferase (GGT, 7-45 U/L), alkaline phosphatase (ALP, 35-100 U /L), and total bilirubin (TB, 3.4-20.5 umol/L) were routinely measured using standard methods. The phenotypic analysis of lymphocytes (CD4+, CD8+, CD3+ T cells) in peripheral blood was performed by a flow cytometry (BD Biosciences; San Diego, CA, USA). We defined abnormal liver damage as any parameter more than the upper limit of normal value (ULN) of ALT (40 U/L), AST(35 U/L), ALP (100 U /L), GGT (45 U/L), TB (20.5 umol/L), LDH (245 U/L).

### Therapeutic strategies

All patients rested in bed and received nutritional support. The basic principle of treatment is symptomatic treatment to maintain the balance of water and electrolyte, stable internal environment. Monitor vital signs and finger oxygen saturation, and give effective oxygen therapy in time. Antiviral therapy can be tried with interferon, lopinavir/litonavir, abidol and dnrunavir. Antibiotics can be used if necessary.

### Statistical Analysis

The continuous measurements were compared by Student’s *t* test or Mann-Whitney U test, which with normal distribution were presented as mean ± standard deviation (SD), while the abnormally distributed measurements as median (Inter Quartile Range), respectively. The categorical variables (shown by percentage) were compared by using Chi-square analysis and Fisher exact test. *P* < 0.05 was determined as with statistically significant differences. Statistical analysis software Graphpad prism 6 was used for all analyses in this study.

### Role of the funding source

The funder of the study had no role in study design, data collection, data analysis, data interpretation, or writing of the report. The corresponding authors had full access to all the data in the study and had final responsibility for the decision to submit for publication.

## Results

### General information of Patients Infected With SARS-CoV-2

Up to January 31, 2020, a total of 148 cases with COVID-19 had been admitted to the Shanghai Public Health Clinical Center, including 73 (49.3%) females and 75 males (50.7%). The average age of the patients was 50 years (36-64). The oldest patient was 88 years old and the youngest was 15 years old. Relative high proportion of the elderly was observed in COVID-19 patients. Patients had clinical manifestations of fever (70.1%), cough (45.3%), expectoration (26.7%) at admission. There were 104 cases of fever, including 47 cases (31.8%) with a temperature between 37.3 and 38 degrees, 45 cases (30.4%) with a temperature between 38.1 and 39 degrees, and 12 cases (8.1%) with a temperature between 39.1 and 40 degrees. The median incubation period in this study was 5 days (3 - 7). Related information of COVID-19 were shown in Table 1.

**Table 1.**
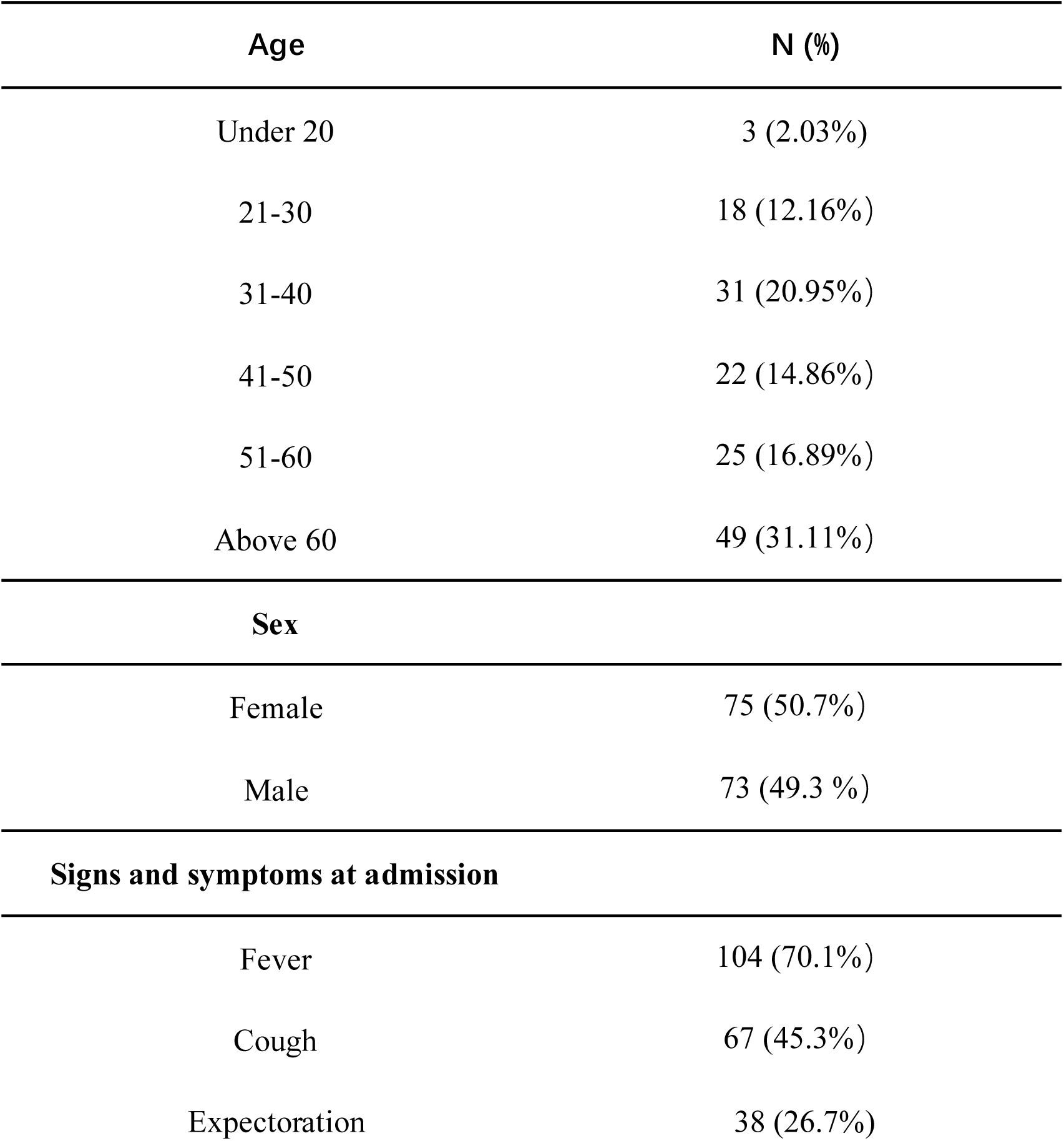
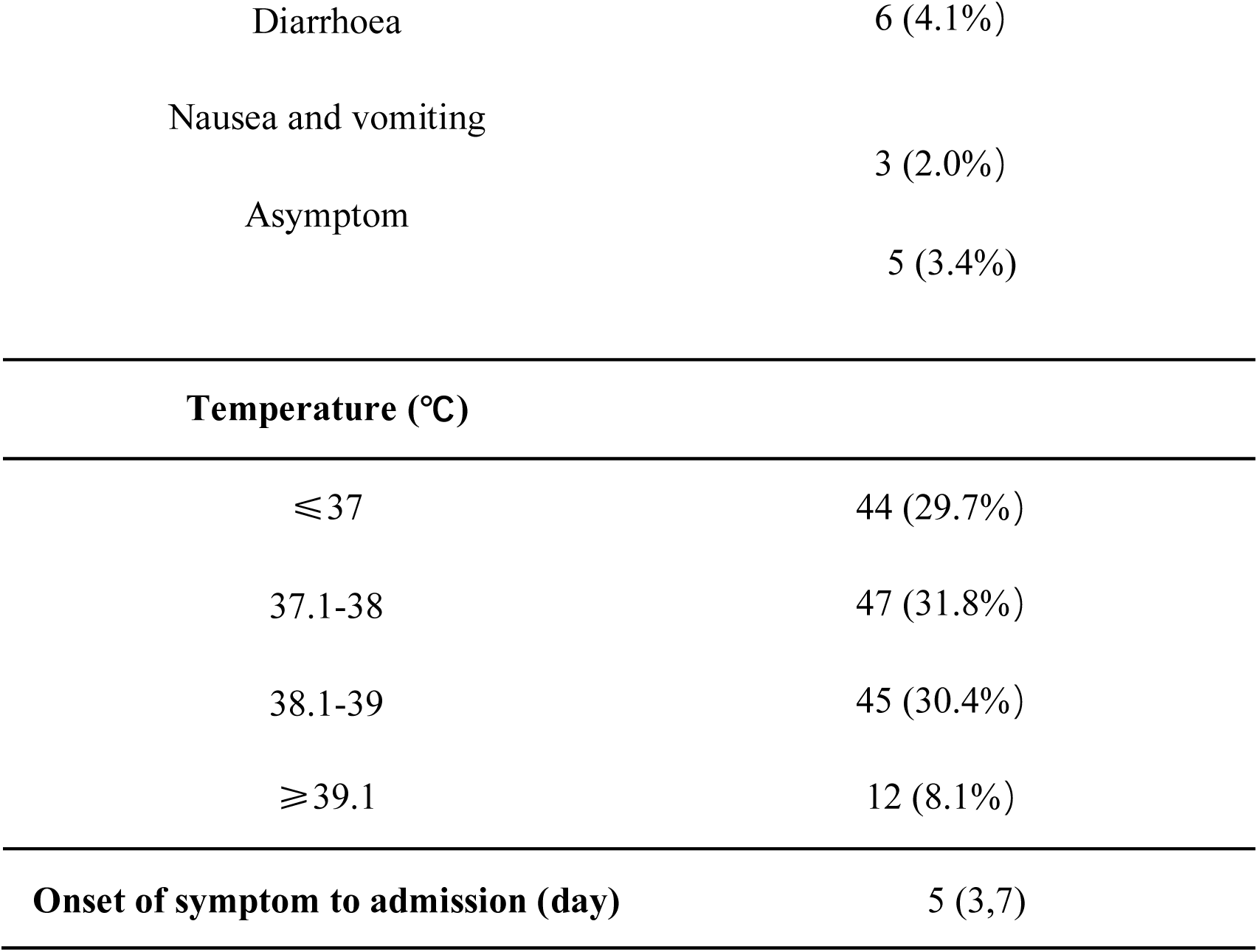
General information of 148 patients infected with SARS-CoV-2 (-Jan 31,2020)

### The proportion and incidence rate of abnormal liver function

There were 75 patients (50.7%) to have abnormal liver function on admission (Figure 1A), including elevated ALT (n = 27, 41-115U/L), AST (n = 32; 37-107U/L), LDH (n = 52, 247-678U/L), GGT (n = 26, 48-159U/L), ALP (n = 6, 102-144U/L), and TB (n = 9, 21-46.6umol/L). As shown in Figure1B, incidence rate of elevated LDH, AST, ALT, GGT, TB, ALP was 35.1%, 21.6%, 18.2%, 17.6%, 6.1%, 4.1% in all patients, respectively.

**Figure 1.**
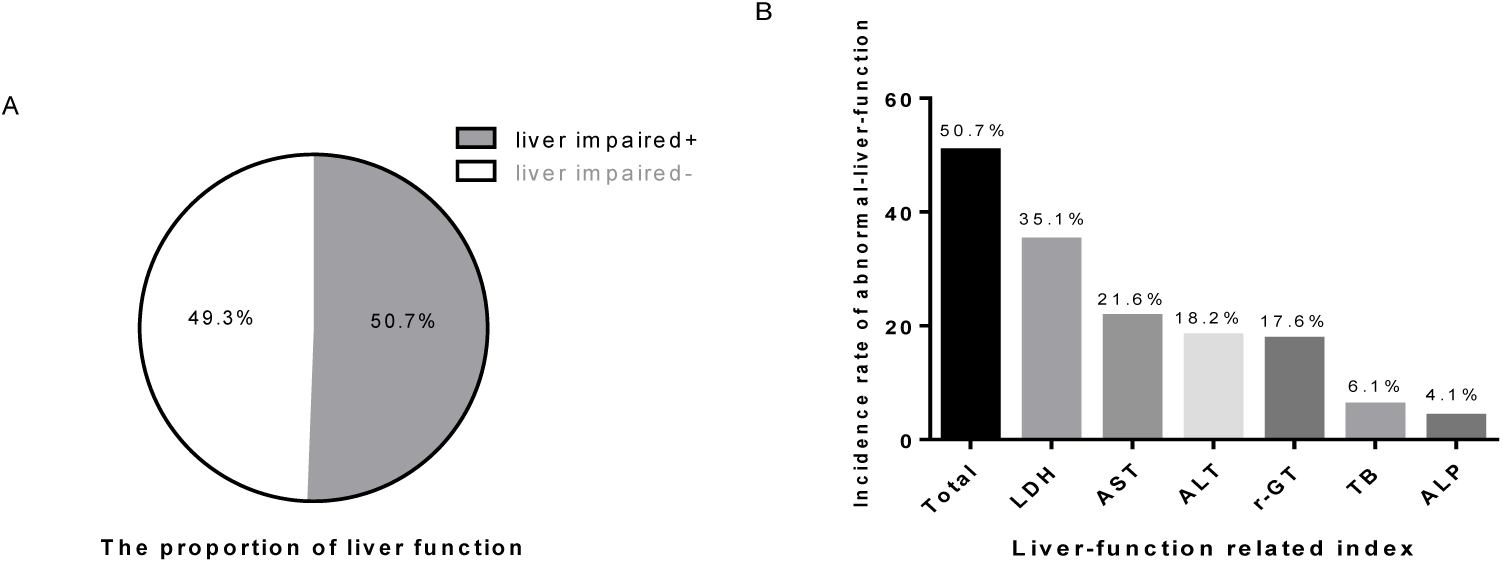
The proportion and incidence rate of normal/abnormal liver function

### Baseline characteristics and medications for patients with abnormal liver function

Compared with patients with normal liver function (n = 73), patients who had elevated liver function index (n = 75) were more likely to have a moderate-high degree fever (44% vs 27.4%; p = 0.035) and significantly present in male (62.67% vs 38.36%; p = 0.005). The medications of COVID-19 patients before admission included antibiotics (Levofloxacin, Azithromycin, Cephalosporin), antiviral drugs (Arbidol, Oseltamivir, Acyclovir)and conventional febrifuge (Ibuprofen). There was no statistical difference in prehospital treatment between two groups (Table 3).

**Table 2.** Baseline characteristics of COVID-19 patients with normal/abnormal liver function

| Liver function | N | Male | Fever | Moderate /high fever | Cough | Expectoration | With other liver diseases |
| --- | --- | --- | --- | --- | --- | --- | --- |
| Abnormal | 75 | 47 (62.67%) | 57(76%) | 33 (44%) | 35 (46.67%) | 17 (22.67%) | 6 |
| Normal | 73 | 28 (38.36%) | 47(64%) | 20 (27.4%) | 32 (43.84%) | 20 (27.34%) | 2 |
| P |  | **0.005** | 0.15 | **0.035** | 0.883 | 0.571 | 0.29 |

**Table 3.**
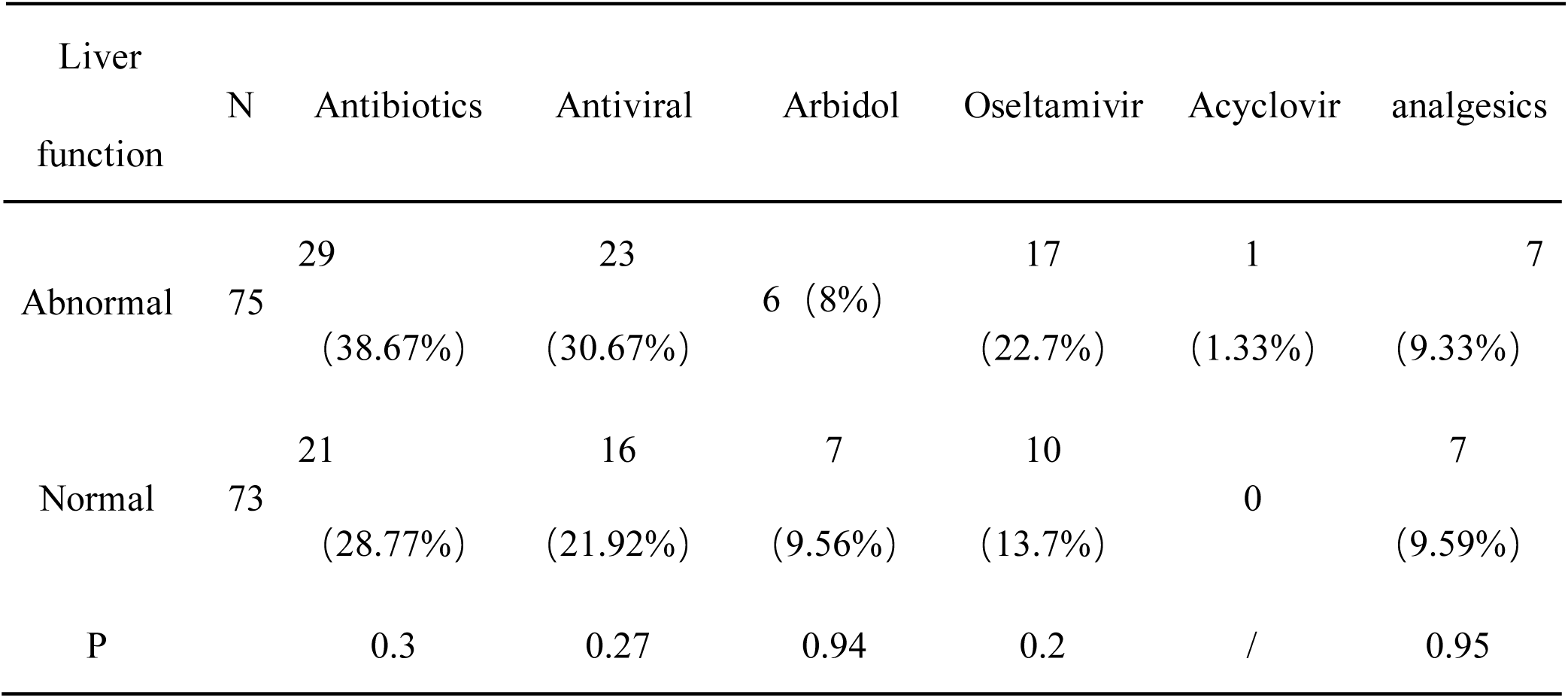
The medications of COVID-19 patients before admission

### Laboratory results of COVID-19 patients

The number of lymphocytes in abnormal liver function group was significantly lower than that in normal liver function group, as shown in Table 4. The numbers of both CD4^+^ and CD8^+^ T cells were lower in abnormal liver function group when compared with those in normal liver function group. Moreover, the levels of PCT and CRP were also observed to be increased in abnormal liver function group than in normal liver function group.

**Table 4.** Laboratory results of COVID-19 patients

| Liver function | Abnormal | Normal | P Value |
| --- | --- | --- | --- |
| WBC | 4.99 (3.743, 5.718) | 4.4 (5.66, 27.5) | 0.3342 |
| lymphocytes | 0.925 (0.635, 1.734) | 1.32 (0.96, 1.57) | **0.0005** |
| PCT | 0.06 (0.03, 0.09) | 0.02 (0.02, 0.04) | **< 0.0001** |
| CRP | 24.7 (12.23, 42.95) | 6.23 (3, 12) | **< 0.0001** |
| ESR | 74 (37.25, 86) | 44.5 (27.5, 82) | 0.1084 |
| CD4+ T | 336 (197, 536.8) | 492 (340, 706) | **< 0.0001** |
| CD8+ T | 201 (126.3, 356.8) | 273 (204, 443) | **< 0.0001** |
| CD3+ T | 551.5 (356.3, 837.3) | 852 (612, 1077) | **< 0.0001** |

### The analysis for medications in COVID-19 patients after admission

We observed that 41 patients with normal liver function had liver injury around 7 (±3) days after admission, with a probability of 56.16%. During the stay in hospitalization, the patients received treatment with antibiotics (Levofloxacin, Meropenem, Moxifloxacin, Cephalosporin), interferon and antiviral drugs (Arbidol, Lopinavir/ritonavir, Dnrunavir). There was no statistical difference in hospital treatment between two groups in terms of antibiotics and interferon used in clinical practice. However, we found that there are more patients with abnormal liver function (56.1%) received treatment with lopinavir/ritonavir compared with those with normal liver function (25%) (p = 0.009)(Table 5).

**Table 5.**
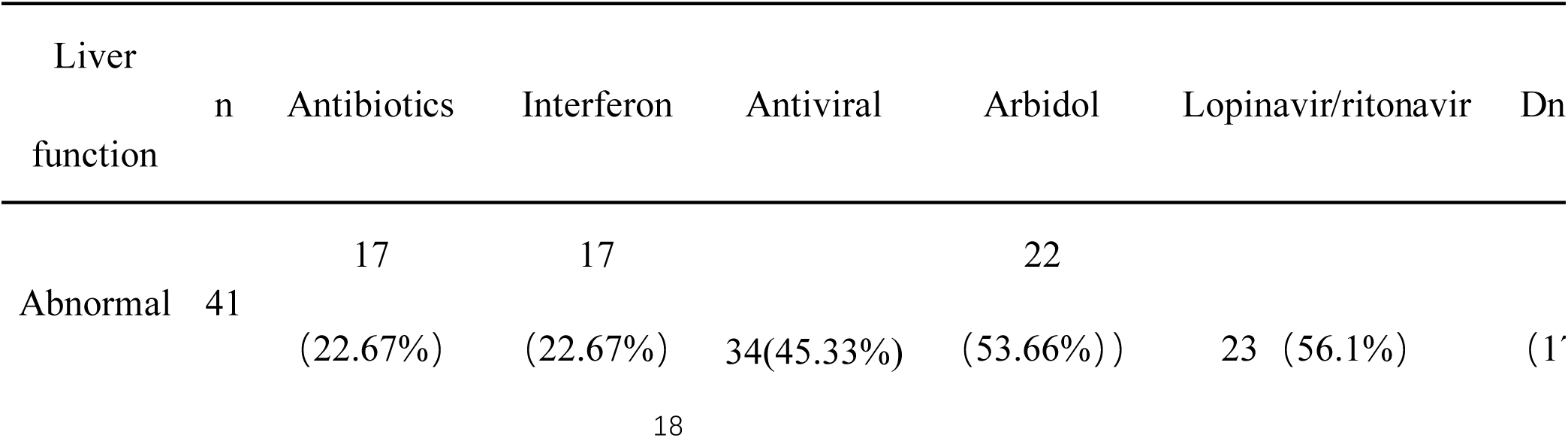
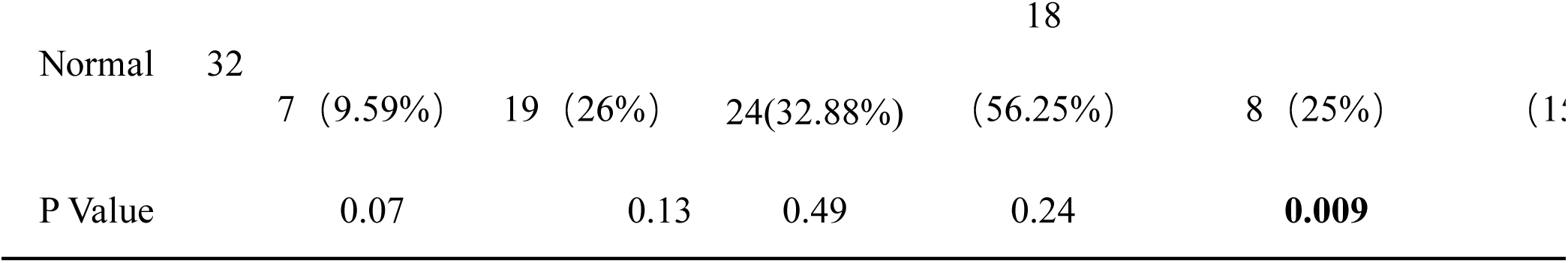
The medications of COVID-19 patients after admission

### The comparison of overall prognosis

Up to February 19, 2020, a total of 93 (62.8%) patients were discharged from hospital, including 44 cases with abnormal liver function before admission, 22 cases emerging abnormal liver function during hospitalization, and 27 cases with normal liver function during the stay in hospital. Of note, we found that liver function related index before admission had little effect on the hospital stays of COVID19 patients, while the emerging abnormal liver function after admission did prolong a length of stay (Table 6).

**Table 6.** Hospital stays of different patients with normal or abnormal liver function

| Timeline | Hospital stays/recovery time days (cases of discharge) |  |  |
| --- | --- | --- | --- |
| Liver function | Before admission | After admission | Total |
| Abnormal | 14.48±4.15 (44) | 16.45±5.19 (22) | 15.14±4.58 (66) |
| Normal | 14.37±5.54 (49) | 12.67±5.31 (27) | 12.67±5.31 (27) |
| P Value | 0.91 | **0.02** | **0.03** |

Hospital stays/recovery time: from admission to discharge(recovery)

## Discussion

Nearly half of the patients in this study are over 50 years old, which is consistent with the previous report.^14^However, there was no significant difference in the prevalence between men and women, while another study including 72314 cases demonstrated that the incidence of COVID-19 is higher in males than that in females. ^15^There are 85% of patients presented with fever, which was similar to 83-98.6% in other reports.^3,8,16^ However, the fever was not an inevitable manifestation of infection. There are even 5 cases of asymptomatic patients in our study, who were actively hospitalized after close contact with the confirmed cases, and then diagnosed with COVID-19. Recently, a case reported an asymptomatic carrier without any symptoms and signs transmitted the coronavirus to other five persons.^17^ Undoubtably, the asymptomatic patients increase the challenge in the prevention of COVID-19 infection. Therefore, we should pay close attention to asymptomatic high-risk groups in clinical diagnosis and treatment.^15^ Further studies are needed to explore the underlying mechanism whereby the asymptomatic patients could exist.

Notably, more than half of patients at admission are presented with abnormal liver functions, which seems to have nothing to do with clinical medication. The proportion of increased ALP was the lowest in patients with liver function injury, and the incidence of increased ALP was also the lowest in all patients admitted to hospital, which is similar to the previous studies.^18^ Although ACE2 is highly expressed in bile duct cells, recent works suggest that SARS-CoV-2 infection did not cause an increase in the marker of bile duct injury, ALP exclusively.^10^ Relatively speaking, elevated markers of liver cells injury (ALT, AST) are more common. Approximately, one in five patients in this study have elevated ALT or AST, which is slightly lower than that in the earlier studies.^3,16^ Moverever, the levels of elevated ALT and AST are generally not high on admission in our study, indicating the COVID-19 – related liver injury is not serious at the initial stage of infection. These findings are consistent with previous research.^8^ Studies have found liver damage is more common in patients with severe pneumonia, which is suspected to be associated with inflammatory factor storm.^3,8^ But it cannot explain the fact that there is also liver damage in mild patients. There are many similarities between the outbreak of SARS-CoV-2 and SARS-CoV. By literature review, we refer to the autopsy analysis about patients died of severe acute respiratory syndrome (SARS).^19,20^ In addition to lungs and immune systems severe damage, fatty degeneration, central lobular necrosis and SARS-CoV were detected in the liver. So there is a reason to believe that SARS-CoV-2 can also attack the human liver.

In addition, it is worth noting that LDH is the most frequently increased in this study (35.1%). According to the existing studies, the proportion of increased LDH in cases from Wuhan is 39.9% - 76%. ^3,8,16^ According to previous research, the levels of LDH in the patients with SARS and MERS are also increased.^21,22^ The level of LDH in the patients is an independent risk factor for SARS.^21^ Due to the poor specificity, LDH may be related to lung, skeletal muscle and myocardium, so it does not fully represent the liver function.^23^ We compared the relationship between LDH level and heat degree in patients, and found that the LDH in patients with moderate/high fever significantly higher in comparation with that in patients with low fever and normal temperature (data not shown). Whether LDH can be used as an early alarm factor for COVID-19 or not needs further analysis.

Although there are also serious liver injuries in some reports, the concomitant chronic liver diseases couldn’t be excluded.^16^ There were 8 cases with chronic hepatitis B or C in our study, but there was no statistical difference in the proportion of chronic liver disease between the abnormal liver function group and normal liver function group.

Because of no effective antiviral drug for COVID-19 available, symptomatic and supportive treatments are rather crucial. Many patients were applied with other antiviral and antipyretic drugs. However, both antiviral drugs and acetaminophen have adverse reactions, such as liver function injury.^24-26^ Previous studies have not further analyzed whether these patients with abnormal liver function are caused by SARS-CoV-2 infection or by the drugs used. In this study, the drugs used by patients before admission are mainly antibacterial drugs (including moxifloxacin, cephalosporins), antiviral drugs (abidol, oseltamivir, acyclovir), and antipyretic drugs with acetaminophen. We analyzed the prehospital medications of the two groups and found that there is no statistical difference between the two groups. Therefore, we speculated that the liver function damage of COVID-19 patients is closely related to virus infection. In addition, the proportion of male patients with liver damage was large, and the specific mechanism was unclear. The body temperature of patients with liver damage was significantly higher than that of the control group, which may be related to the immune response after virus infection.

Patients with liver damage had higher inflammatory indexes, such as elevated CRP and PCT, and the reduced numbers of lymphocyte absolute count and T cells. This is basically consistent with another study and the reduction and functional exhaustion of T cells were found in COVID-19 Patients.^27^ However, the direct relationship between the decrease of T cell count and liver damage remains unclear.

After admission, a part of cases with normal liver function developed liver function injury. Comparing with the medication of these patients during hospitalization, our study found that the application rate of lopinavir/ritonavir in liver damage group was significantly higher than that in patients with normal liver function. What’s more, the length of stay in patients with liver damage after admission was significantly longer than that in cases with normal liver function. In view of another article on antiviral effect in our hospital, lopinavir/ritonavir has no effect on the negative conversion rate of SARS-CoV-2.^28^ For this reason, we tend not to recommend lopinavir/ritonavir as a common clinical treatment drug for COVID-19, even in mild patients with normal liver function. More studies are needed to further evaluate the risks and benefits that lopinavir/ritonavir may bring.

However, there are also deficiencies in our research. This study was retrospective, and some cases had incomplete documentation for the history of present illness. Moreover, all data were collected from a single center at a certain timepoint. So the sample size is relatively limited. Furthermore, the direct evidence of abnormal liver function caused by SARS-CoV-2 has not yet been verified. Further studies are needed to corroborate the pathogenic mechanism.

In conclusion, this is the first time to analyze the clinical features of COVID-19-related liver injury and the relationship between clinical medication and liver damage in SARS-CoV-2-infected patients. SARS-CoV-2 may cause the liver function damage, and the emerging liver injury after admission has some connection with the application of lopinavir/ritonavir and the extend length of hospital stay. The finding is expected to provide some references for the clinical treatment of the current epidemic.

## Data Availability

All data in the manuscript are available

## Notes

### Competing Interest Statement

The authors have declared no competing interest.

### Funding Statement

Shanghai Science and Technology Commission Fund Project 17411969500 and National Science and Technology Major Project 2017ZX10203202-003-007.

